# Self-Collected Anterior Nasal and Saliva Specimens versus Healthcare Worker-Collected Nasopharyngeal Swabs for the Molecular Detection of SARS-CoV-2

**DOI:** 10.1101/2020.07.17.20155754

**Authors:** KE Hanson, AP Barker, DR Hillyard, N Gilmore, JW Barrett, RR Orlandi, SM Shakir

**Affiliations:** Department of Medicine, Division of Infectious Diseases, University of Utah School of Medicine, Salt Lake City, Utah, USA; Department of Pathology, Section of Clinical Microbiology, University of Utah and ARUP Laboratories, Salt Lake City, Utah, USA; University of Utah Hospital and Clinics, Salt Lake City, Utah, USA; Department of Surgery, Division of Otolaryngology – Head and Neck Surgery, University of Utah School of Medicine, Salt Lake City, UT

## Abstract

We prospectively compared healthcare worker-collected nasopharyngeal swabs (NPS) to self-collected anterior nasal swabs (ANS) and straight saliva for the diagnosis of COVID-19 in 354 patients. The positive percent agreement between NPS and ANS or saliva was 86.3% (95% CI: 76.7-92.9) and 93.8% (95% CI: 86.0-97.9), respectively. Negative percent agreement was 99.6% (95% CI: 98-100) for NPS vs. ANS and 97.8% (95% CI: 95.3 – 99.2) for NPS vs. saliva. NPS (n=80) and saliva (n=81) detected more cases than ANS (n=70), but no single specimen type detected all SARS-CoV2 infections.

## Introduction

Rapid and accurate diagnostic tests are essential for controlling the SARS-CoV-2 pandemic. The Centers for Disease Control (CDC) currently recommends collecting and testing an upper respiratory tract specimen for initial SARS-CoV-2 diagnostic testing (1), but the most sensitive specimen type has not been defined. Nasopharyngeal swabs (NPS) have historically been considered the reference method for respiratory virus detection. In addition, anterior nasal swabs (ANS) are used routinely for influenza nucleic acid amplification testing (NAAT). Recurrent shortages of swabs and personal protective equipment (PPE), however, have prompted evaluation of alternatives to NPS including the use of patient self-collected ANS and saliva.

The advantages of ANS and saliva are the minimally invasive nature of sampling and potential for patient self-collection, which may reduce healthcare worker exposure to infectious aerosols. Saliva also has the added benefit being a “swab-free” specimen type known to contain high concentrations of SARS-CoV-2 RNA (2-4). Surprisingly few studies have assessed the performance of self-collected ANS for SARS-CoV-2 testing (5, 6). Small sample sizes and use of selected cases limits the available evidence for ANS. More performance data exists for saliva (7), but published studies vary substantially in the way the specimens were obtained. Many saliva protocols require patients to cough before pooling saliva in their mouth (2, 3, 8), entail avoidance of food, water, or tooth brushing prior to testing (9), and/or rely on RNA stabilization reagents as a part of the collection device. Forced cough, if performed in the presence of a healthcare worker, necessitates the need for PPE and restrictions on eating and drinking are not feasible in all care settings. Furthermore, RNA stabilizers increase the cost of testing, are vulnerable to supply shortages, and may not be compatible with all NAAT chemistries. Larger studies that compare the performance of self-collected ANS and “straight” saliva to NPS for SARS-CoV-2 detection are needed. Therefore, we performed a prospective comparative study to evaluate the performance of self-collected ANS and saliva versus healthcare provider-collected NPS for SARS-CoV-2 diagnostic testing.

## Methods

### Study Subjects

Adult patients presenting to a drive-thru test center with symptoms suggestive of COVID-19 were includes. After obtaining consent, subjects were instructed to swab both nostrils, pool saliva in their mouth, and then repeatedly spit a minimum of 1 mL saliva into a sterile tube in the presence of a healthcare worker. The NPS was collected last in the sampling sequence. The University of Utah Institutional Review Board approved all study procedures.

### Specimen collection

Flocked mini-tip and foam swabs (Puritan Medical Products) were used for the nasopharyngeal and nasal collections. Swabs placed in 3 mL of sterile saline and straight saliva were transported to the clinical laboratory at 4°C. Saliva was diluted 1:1 in ARUP Laboratories universal transport media™ (UTM) prior to testing.

### SARS-CoV-2 detection

All specimens were analyzed using the Hologic Aptima SARS-CoV-2 transcription mediated amplification (TMA) test (Hologic Inc.). Discrepant NAAT results across specimens collected from the same patient triggered repeat testing using the Hologic Panther Fusion (Hologic Inc.), a PCR-based platform, to assess crossing thresholds (Cts) as a surrogate measure of RNA concentration. Cts of ≤ 42 by PCR are considered positive.

### Statistical methods

The standard of care NPS results by TMA were used as the benchmark for assessments of test agreement. GraphPad Quick Calcs software was used to calculate kappa coefficients (k) and proportions (*p* value) by the Chi Square test. Percent positive or negative agreement for categorical variables were calculated in Microsoft Excel using the Analyse-it software package.

## Results

A total of 1104 paired specimens were collected from 368 unique patients between May 29^th^ and June 25^th^, 2020. The average age of study participants was 35 (range 18-75 years), 47% female and 53% male. Saliva samples from 12 patients (3.3%) generated invalid TMA results due to automated sample processing errors or internal control failure and an additional 2 patients did not provide adequate saliva volume for testing. Patients with missing saliva data (n=14) were excluded from the primary analysis.

**Tables 1 and 2** contain the summary of all TMA results. There was near perfect qualitative agreement across sample types (NPS vs. saliva κ=0.912 [95%CI: 0.86-0.96]; NPS vs. ANS κ=0.889 [95%CI: 0.84-0.95]). In all, 66 (18.6%) patients had SARS-CoV-2 detected in all 3 specimens types, 13 (3.7%) in 2 specimens, 7 (2.0%) in 1 specimen, and 268 (75.7%) had completely negative testing. The 7 single specimen positive detections included 2 (28.6%) infections detected by NPS only and 5 (71.4%) by saliva only. Positivity rates were higher for NPS (22.5%; 80/354) and saliva (22.9%; 81/354) compared to ANS (19.7%; 70/354) alone, but this did not reach statistical significance (p = 0.408 for the NPS vs. ANS comparison). The greatest case detection rate combines NPS sampling with saliva (23.6%; 86/354).

**Table 1.**
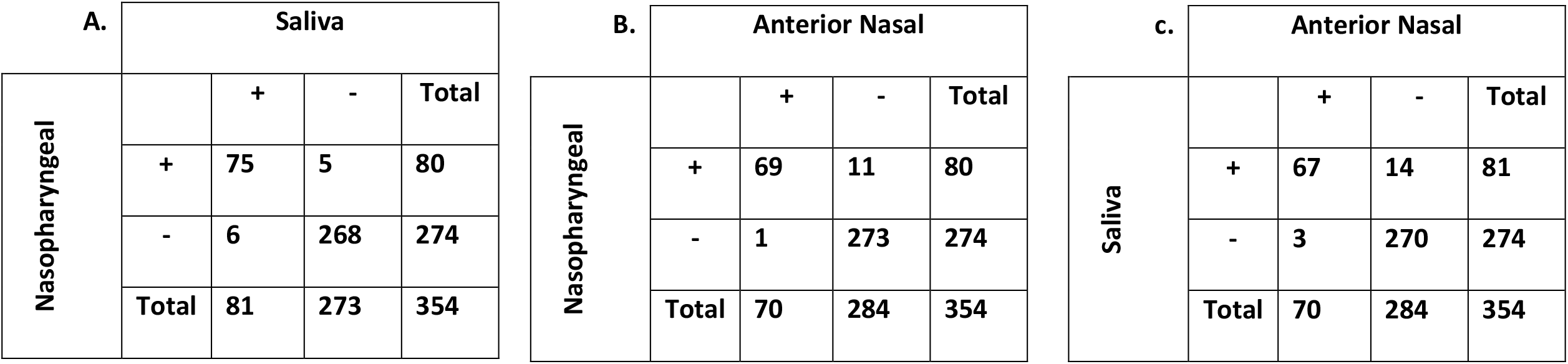
Qualitative Result Comparisons across All Specimen Types.

| A. Saliva |  |  |  |  | B. Anterior Nasal |  |  |  |  | c. Anterior Nasal |  |  |  |  |
| --- | --- | --- | --- | --- | --- | --- | --- | --- | --- | --- | --- | --- | --- | --- |
| Nasopharyngeal |  | + | - | Total | Nasopharyngeal |  | + | - | Total | Saliva |  | + | - | Total |
|  | + | 75 | 5 | 80 |  | + | 69 | 11 | 80 |  | + | 67 | 14 | 81 |
|  | - | 6 | 268 | 274 |  | - | 1 | 273 | 274 |  | - | 3 | 270 | 274 |
|  | Total | 81 | 273 | 354 |  | Total | 70 | 284 | 354 |  | Total | 70 | 284 | 354 |

**Table 2.** Percent Agreement between Nasopharyngeal Swabs and Alternative Specimen Types.

|  | Saliva vs. Nasopharyngeal | Nasal vs. Nasopharyngeal |
| --- | --- | --- |
| Positive Agreement (%) | 93.8 (95% CI: 86.0-97.9) | 86.3 (95% CI: 76.7-92.9) |
| Negative Agreement (%) | 97.8 (95% CI: 95.3-99.2) | 99.6% (95% CI: 98.0 – 100.0) |

Adequate residual sample volume was available for 15 of 19 discrepant specimen sets to perform repeat PCR testing. The average Ct values for NPS positive only or saliva positive only specimens were 27.0 (range 19.7 – 32.7) and 28.2 (range 18.3 – 37.5), respectively. Similar Ct ranges (22.0-35.7) were seen in the NPS positive/ANS negative specimens. For comparison, **Figure 1** displays the distribution of all Ct values generated by the Panther Fusion SARS-Co-V2 assay performed as a part of routine clinical testing in our clinical laboratory since March 2020.

**Figure 1.**
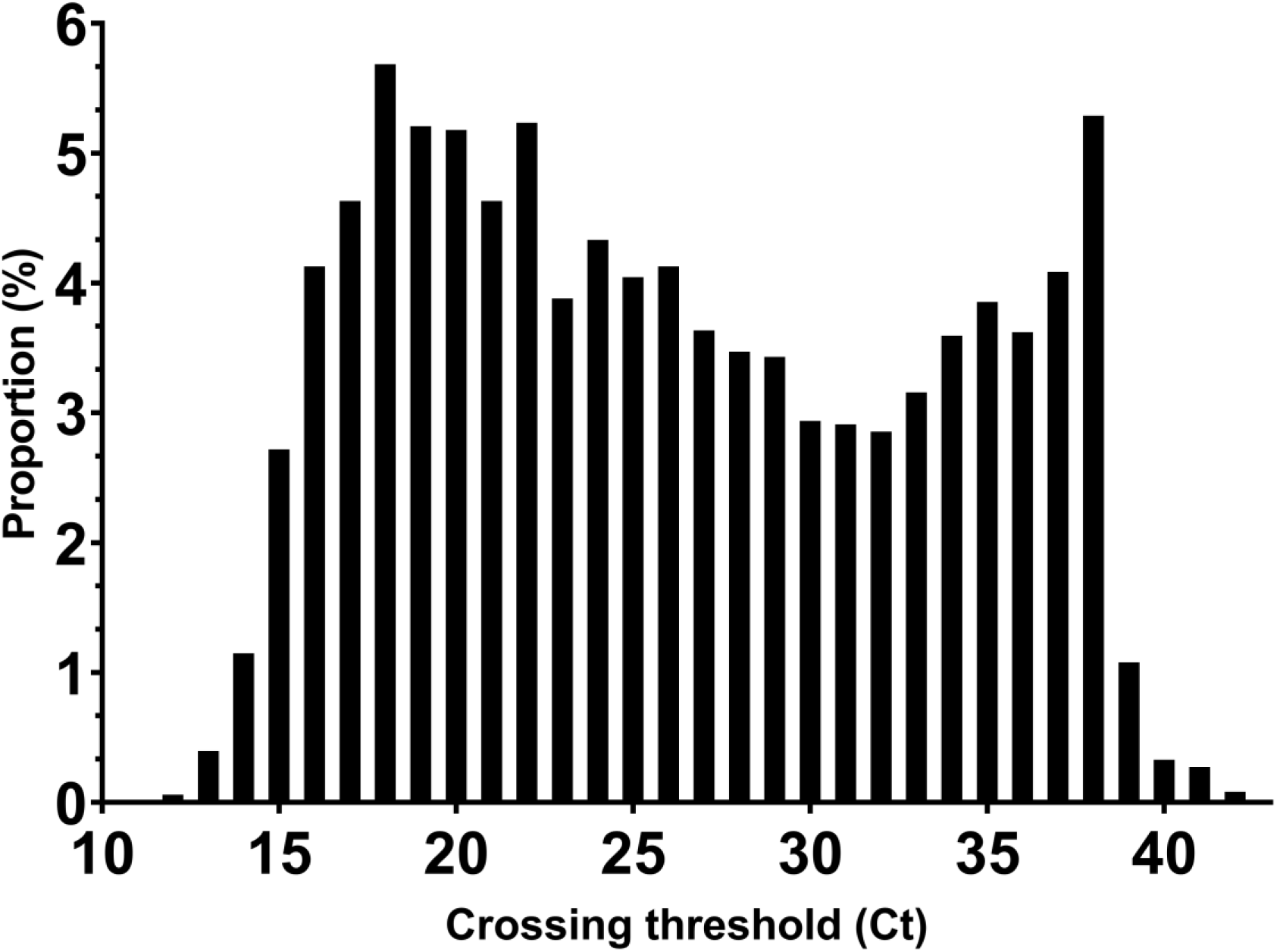
Distribution of PCR Crossing Thresholds (Cts) for all Clinical Specimens.

## Discussion

Sensitive detection of SARS-CoV-2 RNA is critical for patient management decisions, hospital infection prevention, and curbing the ongoing Public Health emergency. The selection and adequate collection of clinical specimens plays an essential role in diagnostic test performance, and this holds true for sensitive NAAT methods. Both the CDC (1) and Infectious Diseases Society of America (10) endorse use of NPS or ANS (either healthcare worker or patient collected) for the diagnosis of COVID-19. However, little data exists comparing the performance of different sample types collected from the same patient, at the same time, and using U.S. Food and Drug Administration (FDA) authorized NAAT platforms.

This study represents one of the largest prospective specimen type comparisons to date and demonstrates excellent agreement between provider-collected NPS and patient-self collected saliva and ANS. The majority (91.9%) of patients with positive results had SARS-CoV-2 nucleic acid detected in at least two specimen types concurrently. NPS and saliva samples had the greatest positivity rates overall. Given that all participants had a strong clinical suspicion for COVID-19, and molecular testing in general has very high specificity, it is likely that the NPS or saliva positive only specimens are true positives; but the lack of an accepted external reference standard precludes calculations of clinical sensitivity and specificity. Even though there was excellent qualitative agreement across specimen types, relying on ANS alone could have missed infection in 10 to 11 patients compared with NPS or saliva, respectively. Missed COVID-19 cases have major clinical implications affecting isolation decisions for symptomatic patients and are a lost opportunity for contact tracing.

No single sample type detected all potential cases and discrepant results were not always explained by high Ct values (i.e. low RNA concentrations near the limit of detection of the test). There are several potential explanations for “false negative” results. First, inadequate swab collection technique is possible. We did not include a host genomic marker to assure presence of respiratory epithelial cells on the swab, nor did we compare self-collection to healthcare provider-collected ANS. Previous respiratory virus studies, however, suggest that self-collected is equivalent to provider-collected ANS (11). Additionally, the level of viral replication in the nasopharynx or posterior oropharynx/salivary glands may vary over the course of infection. We did not collect information on the duration or type of symptoms at the time of specimen collection, which is an additional limitation of the study. Lastly, in an attempt to exclude RNA degradation in straight saliva as a potential explanation for “false negatives”, we performed stability studies at ambient and refrigerated temperatures for up to 5 days and saw no reduced TMA or PCR signal (data not shown).

In conclusion, NPS and saliva were superior to ANS alone for the detection of SARS-CoV-2 in symptomatic patients. These observations, along with other recent reports (9, 12), suggest that straight saliva is an acceptable specimen type for symptomatic patients especially if swab or PPE supplies are limited. However, not all patients could provide adequate volume and saliva is a complex matrix that requires clinical laboratories to validate this specimen type on their respective NAAT platforms. An increased indeterminate or invalid rate (3.3% for saliva vs. 0% for swabs in saline) was observed using the Hologic Panther despite a 1:1 dilution in UTM. This could be related to issues of sample viscosity affecting the automated pipetting and/or internal control inhibition. We did not test whether an additional dilution step would reduce the invalid rate without loss of sensitivity. Combination testing with simultaneous sample collection from multiple anatomic sites may increase SARS-CoV-2 detection rates slightly, but multisite testing could be impractical given current swab and reagent shortages. Requiring two separate NAAT reactions would also increase costs.

## Data Availability

The manuscript contains all relevant data

## Acknowledgements

This study was supported by the ARUP Institute for Clinical and Experimental Pathology.

## References

1. Control CfD. 2020. Interim Guidelines for Collecting, Handling, and Testing Clinical Specimens for COVID-19. Accessed July 14, 2020.

2. To KK, Tsang OT, Chik-Yan Yip C, Chan KH, Wu TC, Chan JMC, Leung WS, Chik TS, Choi CY, Kandamby DH, Lung DC, Tam AR, Poon RW, Fung AY, Hung IF, Cheng VC, Chan JF, Yuen KY. 2020. Consistent detection of 2019 novel coronavirus in saliva. Clin Infect Dis doi:10.1093/cid/ciaa149.

3. To KK, Tsang OT, Leung WS, Tam AR, Wu TC, Lung DC, Yip CC, Cai JP, Chan JM, Chik TS, Lau DP, Choi CY, Chen LL, Chan WM, Chan KH, Ip JD, Ng AC, Poon RW, Luo CT, Cheng VC, Chan JF, Hung IF, Chen Z, Chen H, Yuen KY. 2020. Temporal profiles of viral load in posterior oropharyngeal saliva samples and serum antibody responses during infection by SARS-CoV-2: an observational cohort study. Lancet Infect Dis 20: 565–574.

4. Yoon JG, Yoon J, Song JY, Yoon SY, Lim CS, Seong H, Noh JY, Cheong HJ, Kim WJ. 2020. Clinical Significance of a High SARS-CoV-2 Viral Load in the Saliva. J Korean Med Sci 35: e195.

5. Altamirano J, Govindarajan P, Blomkalns AL, Kushner LE, Stevens BA, Pinsky BA, Maldonado Y. 2020. Assessment of Sensitivity and Specificity of Patient-Collected Lower Nasal Specimens for Sudden Acute Respiratory Syndrome Coronavirus 2 Testing. JAMA Network Open 3: e2012005–e2012005.

6. Noah Kojima FT, Vlad Slepnev, Agatha Bacelar, Laura Deming, Siri Kodeboyina, Jeffrey D Klausner. 2020. Self-Collected Oral Fluid and Nasal Swabs Demonstrate Comparable Sensitivity to Clinician Collected Nasopharyngeal Swabs for Covid-19 Detection. medRxiv doi: https://doi.org/10.1101/2020.04.11.20062372.

7. Zohaib Khurshid SZ, Chaitanya Joshi, Syed Faraz Moin, Muhammad Sohail Zafar, David J Speicher. 2020. Saliva as a non-invasive sample for the detection of SARS-CoV-2: a systematic review. medRxiv doi: https://doi.org/10.1101/2020.05.09.20096354.

8. Chen JH, Yip CC, Poon RW, Chan KH, Cheng VC, Hung IF, Chan JF, Yuen KY, To KK. 2020. Evaluating the use of posterior oropharyngeal saliva in a point-of-care assay for the detection of SARS-CoV-2. Emerg Microbes Infect 9: 1356–1359.

9. Anne Louise Wyllie JF, Arnau Casanovas-Massana, Melissa Campbell, Maria Tokuyama, Pavithra Vijayakumar, Bertie Geng, M. Catherine Muenker, Adam J. Moore, Chantal B. F. Vogels, Mary E. Petrone, Isabel M. Ott, Peiwen Lu, Alice Lu-Culligan, Jonathan Klein, Arvind Venkataraman, Rebecca Earnest, Michael Simonov, Rupak Datta, Ryan Handoko, Nida Naushad, Lorenzo R. Sewanan, Jordan Valdez, Elizabeth B. White, Sarah Lapidus, Chaney C. Kalinich, Xiaodong Jiang, Daniel J. Kim, Eriko Kudo, Melissa Linehan, Tianyang Mao, Miyu Moriyama, Ji Eun Oh, Annsea Park, Julio Silva, Eric Song, Takehiro Takahashi, Manabu Taura, Orr-El Weizman, Patrick Wong, Yexin Yang, Santos Bermejo, Camila Odio, Saad B. Omer, Charles S. Dela Cruz, Shelli Farhadian, Richard A. Martinello, Akiko Iwasaki, Nathan D. Grubaugh, Albert I. Ko. 2020. Saliva is more sensitive for SARS-CoV-2 detection in COVID-19 patients than nasopharyngeal swabs. medRxiv doi: https://doi.org/10.1101/2020.04.16.20067835.

10. Hanson KE, Caliendo AM, Arias CA, Englund JA, Lee MJ, Loeb M, Patel R, El Alayli A, Kalot MA, Falck-Ytter Y, Lavergne V, Morgan RL, Murad MH, Sultan S, Bhimraj A, Mustafa RA. 2020. Infectious Diseases Society of America Guidelines on the Diagnosis of COVID-19. Clin Infect Dis doi:10.1093/cid/ciaa760.

11. Akmatov MK, Gatzemeier A, Schughart K, Pessler F. 2012. Equivalence of self- and staffcollected nasal swabs for the detection of viral respiratory pathogens. PLoS One 7: e48508.

12. Williams E, Bond K, Zhang B, Putland M, Williamson DA. 2020. Saliva as a non-invasive specimen for detection of SARS-CoV-2. J Clin Microbiol doi:10.1128/jcm.00776-20.

